# Assessing the Impact of Mask Usage on COVID-19 Transmission Using a Computer Simulation

**DOI:** 10.1101/2021.06.08.21258593

**Authors:** Roger Lacson, Peter Veldkamp, Conrad Zapanta

## Abstract

**Background:** COVID-19, caused by SARS-CoV-2, is highly contagious and causes substantial morbidity and mortality. Mask usage has been advocated by health professionals to minimize its spread. Thus, it is important to develop a simulation that models SARS-CoV-2 spread in indoor environments to evaluate mask usage effectiveness.

**Methods:** A visual computer simulation was developed with Pygame in Python 3. A virtual indoor supermarket is simulated by a given flow of customers with an initial infection percentage and mask usage percentage who enter, move around, and exit a supermarket with shelves, tables and cashiers to demonstrate a system’s dynamic complexity, i.e. nonlinear interactions of system elements over time. A supermarket was simulated with initial infection rates of 5%, 10%, and 20% and mask use percentages of 0%, 25%, 50% 75%, and 100%. The environmental settings (e.g. shelf number and location) and total customers (N=200) were kept constant.

**Results:** The number of infected customers increased as the percentage of mask usage decreased (p < 0.01). At 5% initial infection, almost no infections were observed at 50% mask usage, with a logarithmic best-fit model (R^2^ = 0.947). At 10% initial infection, the association between mask usage and decrease in number of infections was best fit with a linear model (R^2^ = 0.924). For 20% initial infection, a quadratic model was the best fit (R^2^ = 0.934). While a linear model suggests proportional decreases in infection, the quadratic model suggests more significant reductions in infections at higher rates of mask use (i.e. increasing mask usage from 5% to 10% is less impactful than from 65% to 70%).

**Conclusion:** The results suggest that mask usage has a significant impact on decreasing COVID-19 transmission. Ideally, mask usage should be as high as possible to achieve more significant reductions in COVID-19 infections. Various parameters can be adjusted during simulation as we learn more about SARS-CoV-2 to guide policies for minimizing COVID-19 transmission.

## Introduction

A new type of coronavirus called SARS-CoV-2 was identified in December 2019, which led to the novel COVID-19 infection.^1,2^ This was classified by the World Health Organization (WHO) in January 2020 as a public health emergency of international concern,^3^ and in March 2020 as a pandemic.^4^ COVID-19 has impacted human lives, suffering, and behavior, specifically health-related behavior. The disease has spread to over 100 countries resulting in substantial mortality and morbidity.^4–6^ As the pandemic has progressed throughout the United States, mask use has become a divisive political issue, despite the large body of evidence supporting its effectiveness to reduce SARS-CoV-2 transmission.^7^ With the induction of President Joe Biden as the 46th President of the United States, Biden has signed an executive order to mandate mask use on federal grounds and will advocate for a “100-day mask challenge” to depoliticize mask use.^8^

Thus far, simulations and models to predict SARS-CoV-2 transmission have been developed using the Susceptible-Exposed-Infective-Recovered (SEIR) model showing viral transmission as a function of time using factors such as population, average number of interactions, the basic reproduction constant, and incubation period.^9–11^ However, no simulation has been designed based on physical and spatial interactions between a population and the viral particles emitted by infectious individuals. Furthermore, no simulation attempts to model SARS-CoV-2 transmission in indoor environments, which are enclosed, leading to higher concentrations of airborne droplets.^12^

Several strategies have been proposed to minimize viral transmission, especially indoors. In supermarkets, restricting occupancy, one way traffic along aisles, physical distancing and mask usage have been implemented.^11^ The impact of these strategies, however, have not been rigorously assessed.

The purpose of this study is to evaluate the effectiveness of masks in indoor environments through a computer simulation that uses digital representations of viral particle spread. The infection count per simulation was tracked and modelled to determine how increasing mask usage affects viral transmission.

## Methods

A visual computer simulation was developed with Pygame in Python 3 to model the spread of SARS-CoV-2 in a supermarket. This simulation relied primarily on the physical interactions between customers and viral particles. In the visual simulation, red dots represent approximately 10 viral particles. When red dots come in contact with a customer, a counter is added to an individual if they do not have a mask. If a customer has a mask on, however, they received 0.8 counters as per the approximately 20% filter effectiveness for airborne particles observed for a common cloth mask.^13^ When 28 counters are accumulated, a customer becomes infected. This was based on the proposed infectious dose of SARS-CoV-2 being approximately 280 viral particles.^14^ The respiratory rate of all customers was 20 breaths per minute. Each customer exhaled between 10-20 particles per exhalation, but this number is reduced on average by 80% for customers with masks, as masks filter approximately 80% of exhaled particles.^13^ Red dots travelled in a straight parallel path in front of the customer, the normal movement pattern for particles being exhaled or sneezed.^15^

Customers entered at random intervals with a predetermined maximum occupancy of 60 people (plus 3 store employees). The supermarket has 8 store shelves arranged in parallel, and three cashiers (i.e. store clerks) at the front of the store. Each customer had a random number of shelf locations to visit (between 1 and 6), and once all locations were visited, they approached a cashier. Each customer moved at 2 pixels per frame. An A* path-finding algorithm was implemented to optimize customer movement from location to location without walking through shelves and tables. The A* algorithm is a dynamic search optimization algorithm commonly used in graph theory.^16^ Figure 1 shows a sample screenshot of the simulation.

**Figure 1:**
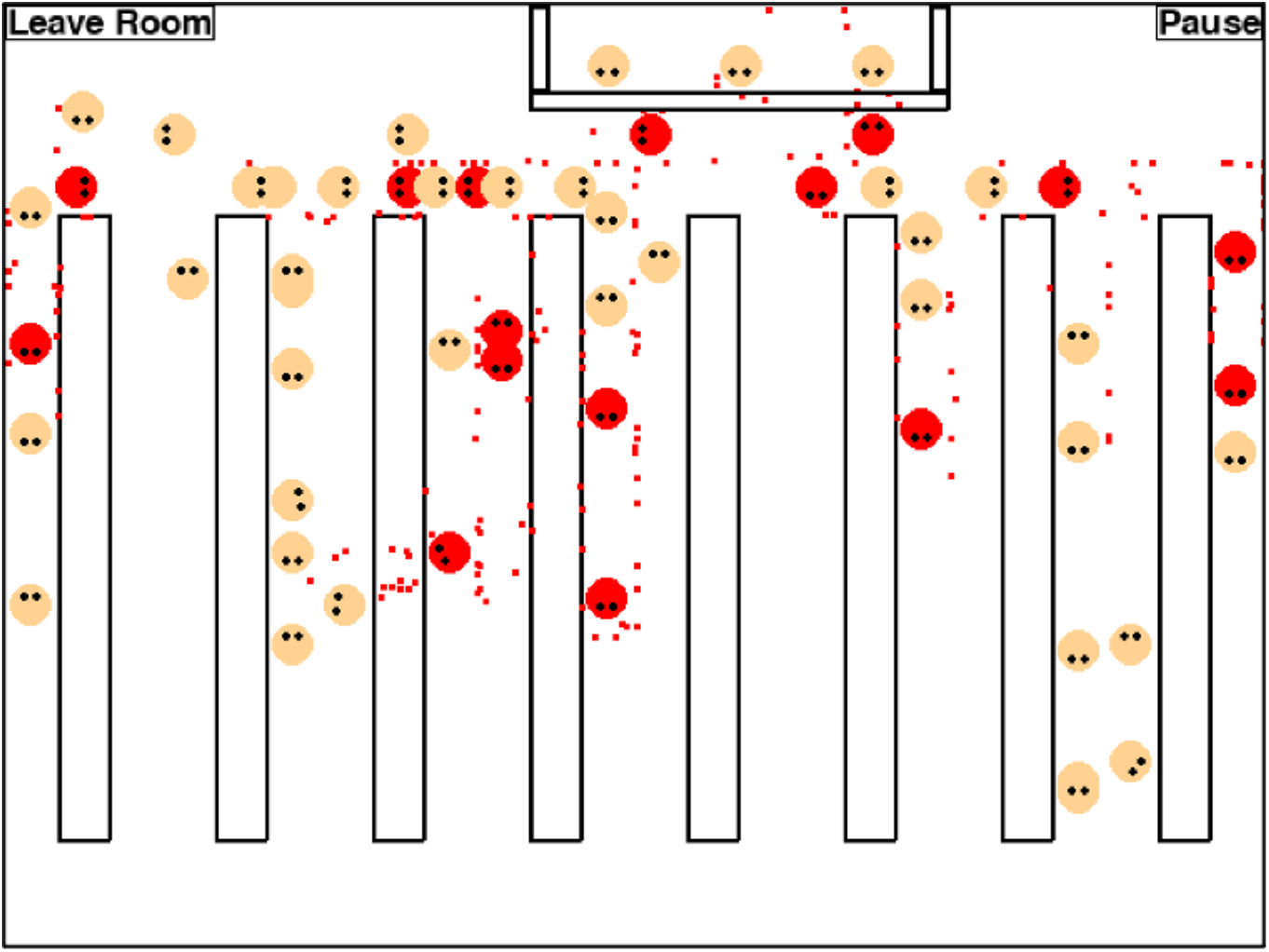
**Sample screenshot of the simulation. Red circles indicate infected individuals that exhale the red dots. Tan circles indicate uninfected individuals. The rectangles represent the shelves or stands in supermarkets. The register is located at the top of the image with three store clerks. The “Leave Room” button for the simulation also serves as the entrance/exit of the supermarket. Note that no distinguishing feature was provided to distinguish mask-users from non-mask-users**.

The virtual indoor supermarket simulates the dynamic flow of customers to demonstrate the system’s complexity, i.e. nonlinear interactions of system elements over time. The inputs in this simulation were mask use, initial infection percentage (percentage of people initially infected), number of store clerks initially infected (out of 3), maximum number of customers allowed in the supermarket, and the percentage of the population at increased risk.

A total of 200 customers were used in each simulation. Mask use varied between 0% (no mask usage), 25%, 50%, 75%, and 100% (full mask usage) to develop a model for mask effectiveness. Initial infection percentages of 5%, 10%, and 20% were tested for each mask condition (i.e. 5% means 10 people of 200 start infected). No store clerks were initially infected. No customers were set at increased risk for contracting COVID-19.

For each mask use percentage and initial infection percentage, a total of 10 simulations were performed. For each simulation, the number of new infections was recorded. The average number of new infections for each condition was plotted against mask use percentage. To fit a plot to the data, various manipulations of mask percentage, including squaring, cubing, exponential, and logarithmic were performed and a linear regression performed. The correlation coefficient R^2^ and the p-value were recorded and a model chosen based on the highest R^2^.

## Results

The number of new infections are included in Table 1 below. There is a strong negative correlation between mask use percentage and number of new infections. This supports the current knowledge suggesting mask use as an effective measure to reduce SARS-CoV-2 transmission.

**Table 1:**
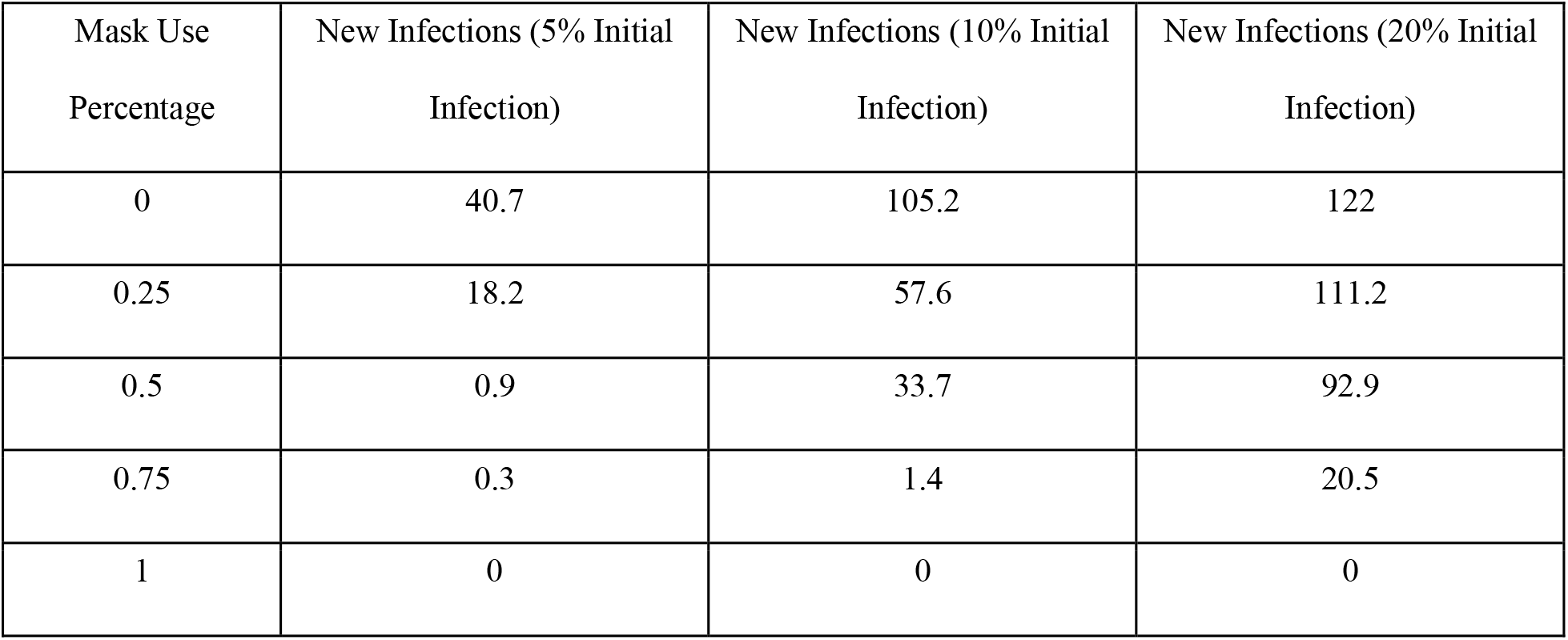
The number of new infections for varied mask use percentages.

To fit a graph to the above data, the correlation coefficient R^2^ for each manipulation of mask use percentage is included in Table 2. The model with best fit (highest correlation coefficient) was logarithmic for 5% initial infection (R^2^ = 0.947), linear for 10% initial infection (R^2^ = 0.924) and quadratic for 20% initial infection (R^2^ = 0.934). There was a statistically significant association between mask usage and number of new infections (p < 0.01). The logarithmic, linear, and quadratic models for 5%, 10%, and 20% initial infection respectively, are graphed in Figure 2.

**Table 2:**
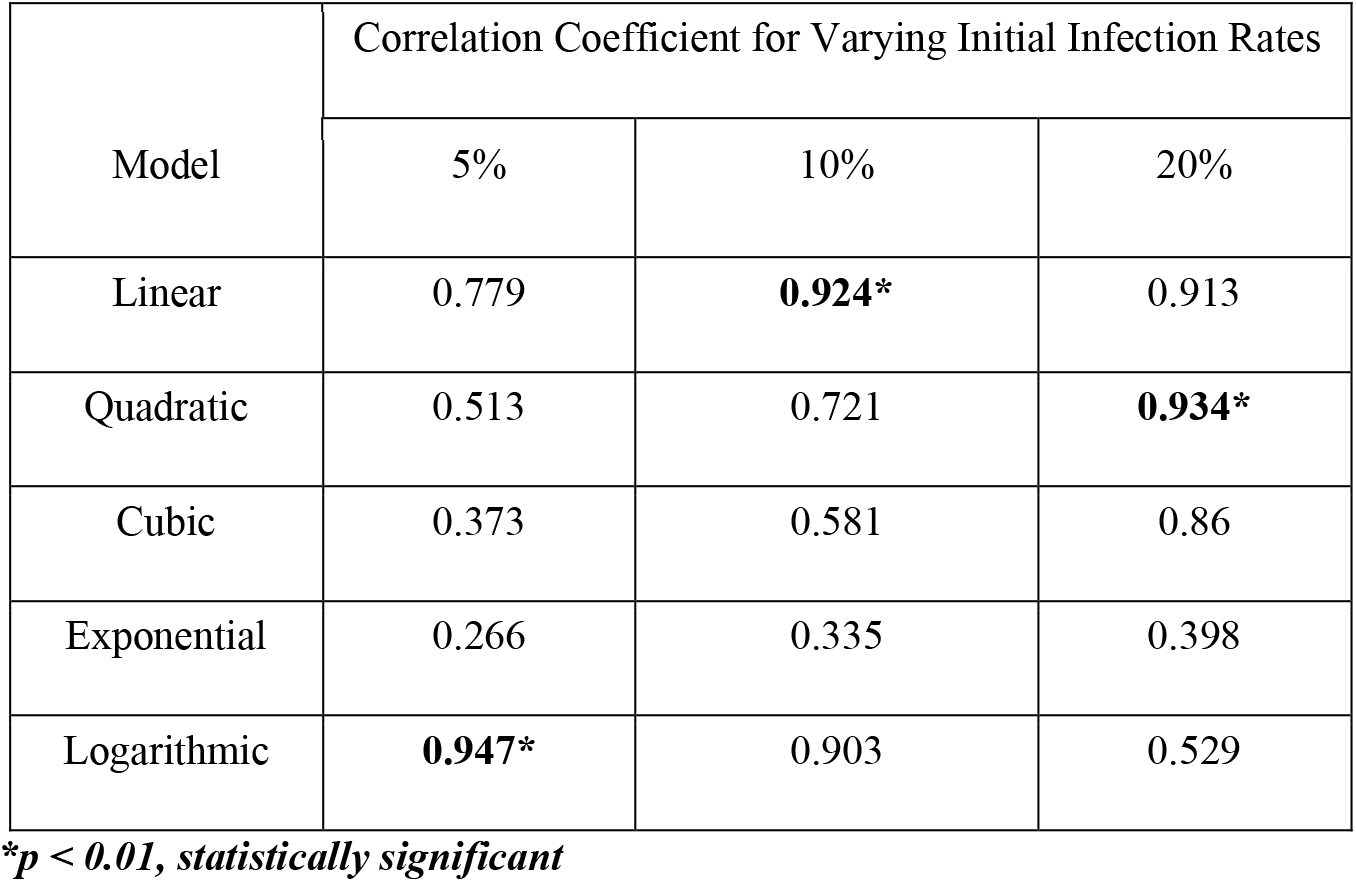
Correlation Coefficient and p-value for different models using linear regression and manipulation of the mask use percentage.

| Model | Correlation Coefficient for Varying Initial Infection Rates |  |  |
| --- | --- | --- | --- |
|  | 5% | 10% | 20% |
| Linear | 0.779 | **0.924\*** | 0.913 |
| Quadratic | 0.513 | 0.721 | **0.934\*** |
| Cubic | 0.373 | 0.581 | 0.86 |
| Exponential | 0.266 | 0.335 | 0.398 |
| Logarithmic | **0.947\*** | 0.903 | 0.529 |

*\* $p < 0.01$ , statistically significant*

**Figure 2a:**
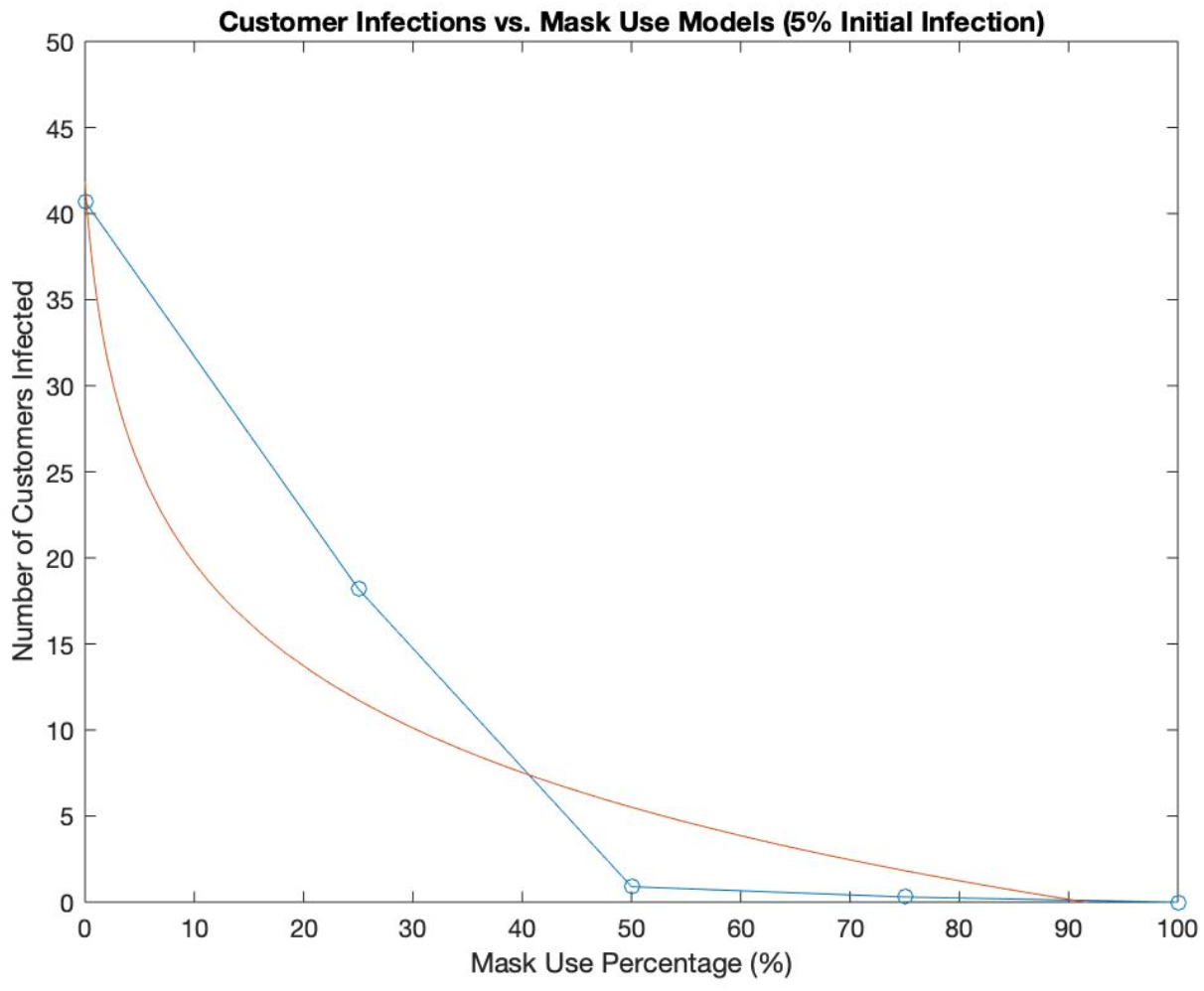
Graph of logarithmic model over the raw data for 5% initial infection.

**Figure 2b:**
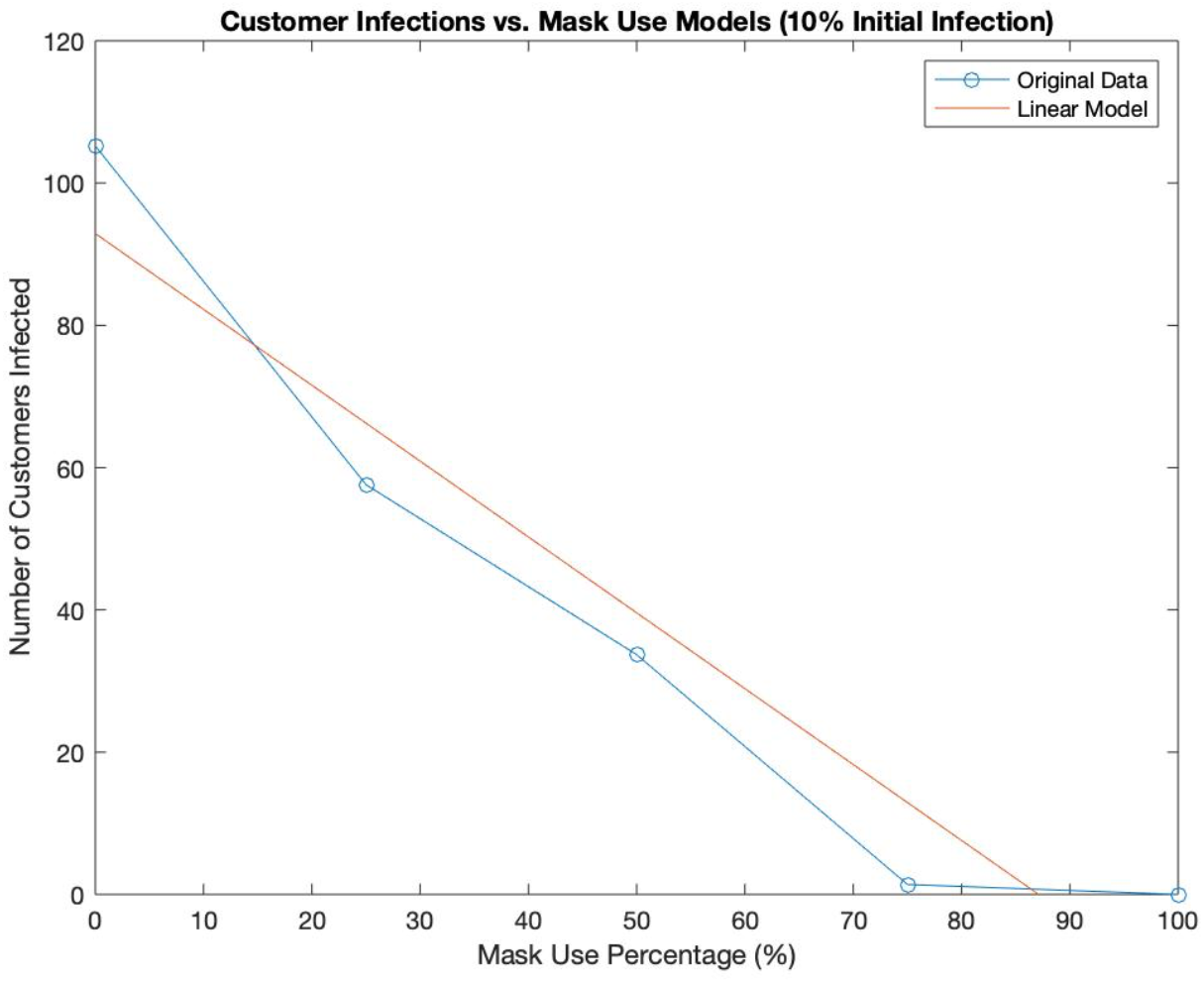
Graph of linear model over the raw data for 10% initial infection.

**Figure 2c:**
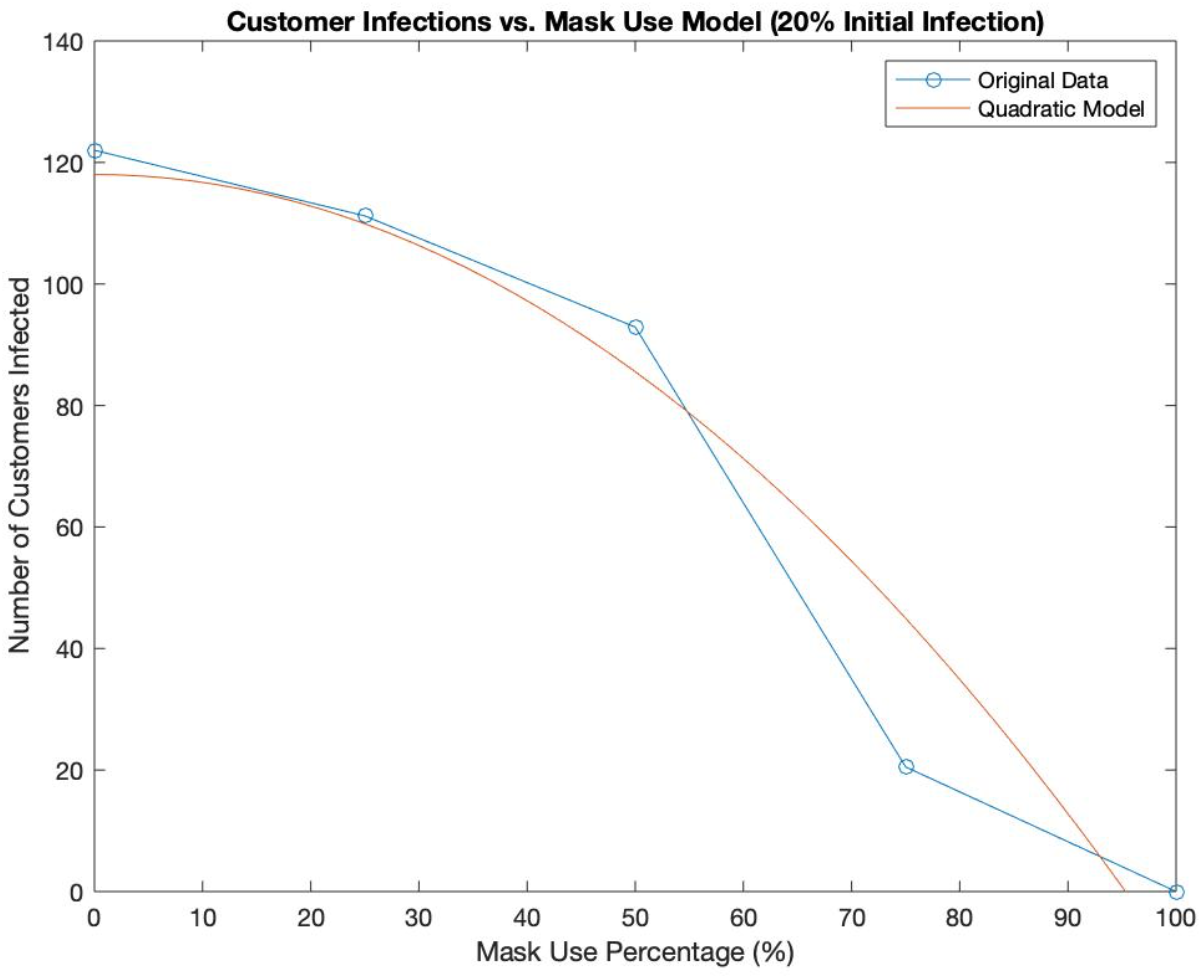
Graph of quadratic model over the raw data for 20% initial infection.

## Discussion

As mask use increased, the number of customer infections decreased, almost entirely diminished at 50% mask use at 5% initial infection, and according to a logarithmic model at 5% initial infection, a linear model at 10% initial infection, and a quadratic model at 20% initial infection. The negative association between mask use and viral transmission was to be expected given that mask use both provides slight protection to users and an even larger protection to those around an infected individual. Furthermore, this conclusion agrees with the common idea that mask use is an effective measure to reduce SARS-CoV-2 transmission, even in indoor environments.

In previous studies,^17–19^ mask usage was an independent factor that was associated with an increased odds of transmission control with an odds ratio of 3.5.^19^ Based on the significant reduction in infections at 50% mask usage for 5% initial infection and the linear model for 10% initial infection, mask use should be encouraged due to the direct effect it has on reducing new infections. However, based on the quadratic model for 20% initial infection, it is important to note that more significant reductions in infections can be observed at higher rates of mask use (i.e. increasing mask usage from 5% to 10% is less impactful than from 65% to 70%). This suggests that in order to have a 50% reduction in infections, the required mask usage should be higher. Based on the quadratic model in Figure 3, this happens at approximately 66% mask usage with more significant decreases in viral transmission occurring at these higher mask use percentages.

### Future Works

While this simulation provides a solid foundation for modelling the spread of SARS-CoV-2 in indoor environments, there are many additional factors that could be added to further improve the accuracy of this simulation. One such factor is re-circulation which could circulate viral particles in indoor environments. This could lead to transmission from infected individuals to others outside the range of exhalation.^12^ Another factor to include is the incubation period. Under this simulation, infected individuals immediately begin exhaling viral particles when there is an incubation period before such spread occurs significantly.^20^ A third factor to consider is adjusting the parameters for the more infectious strains of SARS-CoV-2 such as the B.1.1.7 variant from the United Kingdom^21^ and the B.1.351 variant from South Africa.^22^ A final consideration to improve the accuracy of the simulation is details specifically related to supermarket environments or other indoor environments in general. Such factors for a supermarket include its size and how the shelves are arranged. Additionally, other environments, such as classrooms or sports arenas could be modelled using this simulation, given the proper parameters and characteristics.

## Conclusion

Under simulated conditions, mask usage was demonstrated to be effective in reducing viral spread in indoor environments. Ideally, mask usage should be as high as possible to achieve more significant reductions in COVID-19 infections. Additionally, in order to improve the accuracy of this simulation, adjustments could be made to account for ventilation, incubation period, and specific environment factors. Once the adjustments are included, further simulation can be run to test the effectiveness of other preventative measures, both in isolation and in combination with other measures.

## Data Availability

The study consisted of computer simulated data which will not be provided.

